# Differential shared genetic influences on anxiety with problematic alcohol use compared to alcohol consumption

**DOI:** 10.1101/2020.08.21.20179374

**Authors:** Sarah M. C. Colbert, Scott A. Funkhouser, Emma C. Johnson, Charles Hoeffer, Marissa A. Ehringer, Luke M. Evans

## Abstract

Anxiety disorders and alcohol use disorders are common psychiatric illnesses. Comorbidity of the two disorders can have a tremendous effect on treatment of one or both disorders, as well as an individual’s social, economic, and physical well-being. We estimated genome-wide genetic correlations between anxiety and alcohol use traits using linkage disequilibrium score regression (LDSC) and found strong and positive correlations of anxiety with problematic alcohol use (PAU), but not with most alcohol consumption (AC) measures. We observed strong, positive between-sex genetic correlations for all traits, but found suggestive evidence that the genetic correlation between alcohol use and anxiety might differ between males and females. Estimates of local genetic covariance demonstrated divergent genetic covariance profiles of PAU and AC with anxiety phenotypes and localized 12 specific genomic regions that likely contribute to both anxiety and alcohol use. Finally, partitioning the genetic covariance among functional annotations also identified the amygdala, caudate basal ganglia and frontal cortex as contributing significantly to positive genetic covariance between anxiety and PAU phenotypes. This study serves as a framework for an approach to be used in future analyses of the genetics of comorbid disorders.

## Introduction

Anxiety disorders and alcohol use disorders are common psychiatric disorders that have a high rate of comorbidity, co-occurring at a rate two to three times more likely than would be expected if independent [1–4]. There is strong evidence that shared genetic factors contribute to this co-occurrence [5–7]. However, the specific underlying genetic mechanisms of this comorbidity are generally unknown.

Previous research has found that combinations of different anxiety disorders and alcohol use traits show varying rates of co-occurrence. Specifically, anxiety disorders are more likely to co-occur with problematic alcohol use (PAU) compared to alcohol consumption (AC) [2–4, 6, 8, 9]. Recent work demonstrated that when alcohol use, determined by the Alcohol Use Disorders Identification Test (AUDIT) [10], is partitioned into a problem use score (AUDIT-P score) and consumption score (AUDIT-C score), these phenotypes are genetically separable [11]. PAU and AC have both shared and distinct underlying risk loci, and distinct genetic correlations with psychiatric traits, such as seeking out professional help for “nerves, anxiety or depression,” or the worry subcluster of neuroticism, but these studies did not include comparisons specifically with anxiety disorders [11,12]. The overlapping genetic architecture between alcohol use traits and anxiety has not been well-characterized but could lead to a better understanding of the neurobiology and comorbidity of the disorders.

Various methods have been developed to partition genome-wide parameters— such as heritability and genetic covariance—along functional annotations and linkage-disequilibrium (LD) blocks [13–16]. These partitioned estimates can be used to identify potentially pleiotropic effects. This knowledge can enable the development of plausible hypotheses about where in the genome and in which specific tissues shared biological mechanisms may exist. Such biologically relevant information will be critical to facilitate direction for functional follow-up studies, for example using mouse models.

Here, we examine the shared genetic architecture of alcohol use and anxiety disorders. First, we evaluate genetic correlations at a genome-wide scale (both sex- and non sex-stratified). Second, we localize genomic regions underlying these genome-wide correlations. Third, we apply partitioned genetic covariance analyses to annotate specific functional and brain regions involved. Importantly, we distinguish between PAU and AC, and find these two traits have different genetic relationships with anxiety. The framework applied here using GWAS summary statistics and bioinformatics tools can be broadly applied to investigate the shared genetic etiology of pairs of complex traits.

## Methods

### Preregistration

We preregistered our analysis (https://osf.io/kuqgf). All preregistered analyses are reported below or in the supplementary information. We deviated from our preregistered analysis in the specific software package used for estimating local genetic covariance (SUPERGNOVA [16]) because of statistical power and substantial sample overlap between the summary statistics. After performing these preregistered analyses, we expanded the scope of the study to include sex-stratified analyses and brain-specific expression partitioning of genetic covariance (in addition to general, broad tissue-specific annotations), fully described below and in the supplementary information.

### Study samples and phenotypes

We utilized published, publicly-available summary statistics of eight phenotypes from five separate studies (Table 1). All summary statistics were derived from analyses which limited samples to individuals of European ancestry.

**Table 1.** Description of traits: References and total sample sizes for phenotypes used in the analyses.

| *Phenotype* | *Reference* | *N* |
| --- | --- | --- |
| Anxiety meta-analysis | Purves et al., 2019 Mol Psych | 114 091 |
| Anxiety NeuroGenetics STudy | Otowa et al., 2016 Mol Psych | 17 310 |
| UK Biobank Lifetime Anxiety | Purves et al., 2019 Mol Psych | 83 566 |
| Alcohol Dependence | Walters et al., 2018 Nat Neuro | 38 686 |
| AUDIT-P score | Sanchez-Roige et al., 2019 Am J Psych | 121 604 |
| AUDIT-C score | Sanchez-Roige et al., 2019 Am J Psych | 121 604 |
| AUDIT-T score | Sanchez-Roige et al., 2019 Am J Psych | 121 604 |
| Drinks per Week | Liu et al., 2019 Nat Genet | 537 352 |

We drew anxiety phenotype summary statistics from two independent studies and a meta-analysis of them by Purves et al. [17]. Purves et al. meta-analyzed the published summary statistics of the Anxiety NeuroGenetics STudy (ANGST) case-control study [18] and a GWAS of UK Biobank lifetime anxiety disorder data [17]. Both used DSM-based diagnoses of the five core anxiety disorders (generalized anxiety disorder (GAD), panic disorder, social phobia, agoraphobia and specific phobia) to define cases, but differed in their diagnostic criteria. The UKB lifetime anxiety disorder study considered a self-report of a lifetime professional diagnosis of any core anxiety disorder (n cases = 21 108, with 62% of cases being reports of GAD) as well as a probable diagnosis of GAD using questions from the CIDI-SF [19] questionnaire (n cases = 4 345) to be a DSM-based diagnosis (total n cases = 25 453 and total n cases and controls = 83 566). The ANGST study used standardized assessment instruments modified from the CIDI to determine a lifetime diagnosis of any core anxiety disorder in subjects (n cases = 7 016 and total n = 17 310). The assessment instruments used by the six samples in the ANGST study differed to some degree. For example, one cohort included individuals with self-reported panic attacks as cases (despite being subdiagnostic), while all other cohorts required panic disorder diagnoses for cases. We utilized all three summary statistic datasets to assess the role of underlying diagnostic criteria.

The alcohol use traits included both PAU and AC traits. First, we included the drinks per week (DrnkWk) phenotype of the GWAS & Sequencing Consortium of Alcohol and Nicotine (GSCAN) [20], which was the reported number of alcoholic drinks an individual consumed per week (N = 537 352). The AUDIT results reported by Sanchez-Roige et al. [11] are based on ten questions from UKB and 23andMe data, although only the UKB summary statistics were publicly released (N = 121 604). These ten questions together make the AUDIT-Total (AUDIT-T) phenotype. Three of these AUDIT questions refer to general consumption habits, such as quantity and frequency, and together make the AUDIT-Consumption (AUDIT-C) phenotype, while the remaining seven referring to problematic consequences of drinking, such as alcohol-induced blackouts, comprise the AUDIT-Problem Use (AUDIT-P) phenotype. Summary statistics of a GWAS meta-analysis of alcohol dependence, defined by clinician ratings or semi-structured interviews, from Walters et al. [21] was the final alcohol use phenotype (N = 38 686).

### Analyses

#### Genome-Wide Genetic Correlations

We used cross-trait LD score regression (LDSC) [22] to estimate genome-wide genetic correlations (r_g_) among pairs of alcohol use traits and anxiety disorders. We also used LDSC to estimate sex-stratified genetic correlations among pairs of alcohol use disorders and anxiety. Because the published summary statistics were not sex-stratified, we performed the sex-stratified GWAS using UK Biobank data. Sex-stratified GWAS and phenotype definitions are detailed in the supplementary information (Supplementary Methods, Table S1, Figure S1).

#### Local Genetic Covariance

Next, to identify specific regions of the genome underlying the genome-wide genetic relationships between traits, we estimated local genetic correlations between all trait pairs in approximately independent LD blocks using SUPERGNOVA [16]. All 2 353 local genomic regions provided by Zhang et al. [16] were used in the analysis. SUPERGNOVA is robust to sample overlap and therefore no correction was needed. Local genetic covariance significance was based on a Bonferroni-corrected threshold of p< 0.05/(number of trait pairs x number of annotations)< 7.60×10^−7^.

#### Partitioned Genetic Covariance

Finally, we stratified genetic covariance between all trait pairs by genome functionality, tissue type and brain region annotations using the genetic covariance analyzer (GNOVA) [23]. Using GenoCanyon annotations, we were able to partition the genome into two categories, functional and nonfunctional[24]. To partition the genome into seven broad tissue categories, we used GenoSkyline-Plus annotations [25]. In addition to these annotation files provided by Lu *et al*. [23] we also used annotations of brain region-specific expression, which were derived from GTEx [26] brain region annotations provided by Finucane et al. [14]. Sample overlap correction, implemented within GNOVA, was used on the 24 trait pairs known to have substantial sample overlap, while correction was not applied to the four trait pairs with no known overlap (Table S2). Studies were considered to have overlap if the same cohort was used in both of the studies. For example, UK Biobank data were included in the summary statistics from both the AUDIT study and anxiety meta-analysis, therefore substantial sample overlap was assumed. To determine significance of estimates of stratified genetic covariance we used a Bonferroni-corrected threshold of p< 0.05/(number of trait pairs x number of annotations). We also assessed the shared genetic components of these traits stratified by functional annotations using partitioned heritability analysis and correlation of partitioned heritability among traits (Supplementary Methods). As noted in the Supplementary Methods, estimates of correlation of partitioned heritability are likely sample size-dependent, and we include them here for completeness as they were pre-registered analyses, but limit our interpretation.

### Results

#### Genome-Wide Genetic Correlations

We observed significant, positive genetic correlations between all anxiety phenotypes. Similarly, all pairwise analyses between alcohol use traits demonstrated significant, positive genetic correlations (Figure 1, Table S3). While all alcohol use pairwise genetic correlations were significantly positive, the patterns clearly differentiated into two blocks. The first block related to AC, with the highest correlations between DrnkWk, AUDIT-T and AUDIT-C (r_g_≥0.83). At a false discovery rate of 5%, these phenotypes had significant, but slightly weaker genetic correlations with alcohol dependence and AUDIT-P (r_g_≤0.81). Genetic correlations with alcohol dependence were the lowest of all the alcohol-use phenotypes, although notably the correlation with AUDIT-P was relatively strong, with these two traits making up the second block relating to PAU.

**Figure 1.**
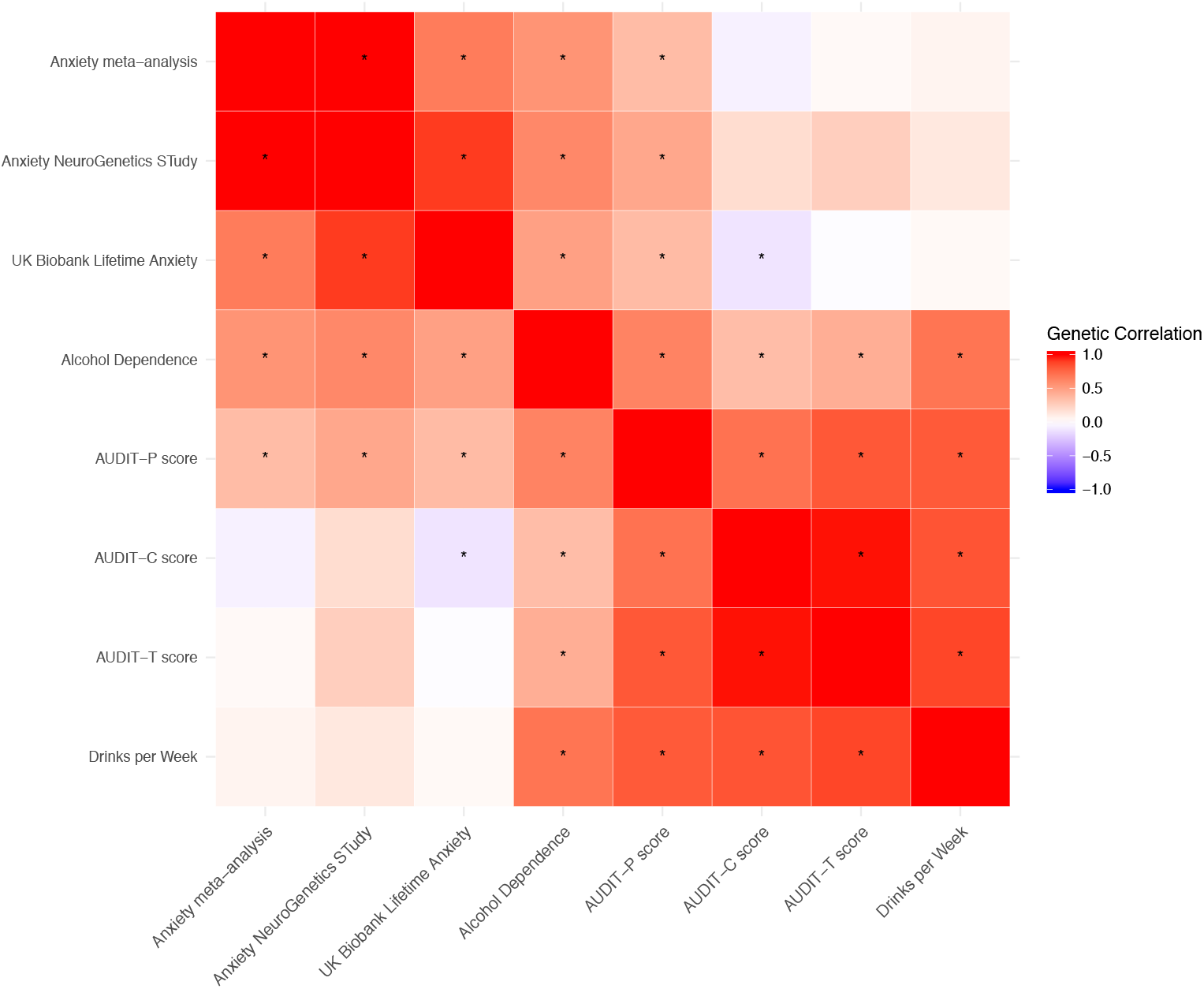
Genome-wide genetic correlations among eight traits using LDSC. Asterisks indicate significant (FDR < 5%) genetic correlations. See Table S3 for complete numerical results.

We found that genome-wide genetic correlations between anxiety and alcohol phenotypes also separated into two groups. All anxiety phenotypes showed significant positive genetic correlation with AUDIT-P and alcohol dependence (r_g_≥0.35). However, the anxiety phenotypes were uncorrelated with AUDIT-T, AUDIT-C and DrnkWk, indicating AC was not genetically related to anxiety. One exception was that we observed a significant, weak negative genetic correlation between AUDIT-C and UKB anxiety (r_g_ = –0.12).

#### Sex-stratified Genome-Wide Genetic Correlations

Genetic correlation between males and females was significant and substantial for all six phenotypes examined (r_g_≥0.75) (Figure S2a, Table S4). We also calculated genetic correlations among traits within each sex (Figure S2b, Table S5). Genetic correlations between alcohol use traits did not differ dramatically between males and females, with all pairs being significantly, positively correlated (r_g,males_ = 0.64–0.95; rg,females = 0.54–0.95). Genetic correlations between anxiety traits differed somewhat between males and females, with males showing higher genetic correlations amongst anxiety traits (r_g_≥0.97) than females (r_g_≥0.75).

We also found sex differences for genetic correlations estimated between anxiety and alcohol use traits (Figure S2b, Table S5). We identified no significant genetic correlations between anxiety and alcohol use disorders in males. Conversely, we identified three, positive, significant genetic correlations between anxiety and alcohol use disorders in females (FDR < 5%). AUDIT-P score was significantly correlated with DSM-V-like GAD (r_g_ = 0.31) and any anxiety disorder (r_g_ = 0.59) in females. AUDIT-T was also significantly correlated with any anxiety disorder (r_g_ = 0.52).

#### Local Genetic Covariance

We identified a total of 47 independent loci with significant (p< 7.60×10^−7^) local genetic covariance between pairs of traits (Table S6). A majority of the regions in which we identified significant local covariances comprised pairs of alcohol use or pairs of anxiety traits (Figures S3-S9). However, we did identify 12 regions with significant local genetic covariances between anxiety and any alcohol use phenotype (Table S6). Three genomic regions showed significant positive local genetic covariance between PAU and anxiety phenotypes (Figure 2a), including two (chr9:10 879 188–11 616 822; chr5:102 686 157–104 542 488) that showed no significant local genetic covariance with AC. Only one region (chr3:85 093 629–86 734 415) had significant negative covariance between PAU and anxiety (Figure 2a).

**Figure 2:**
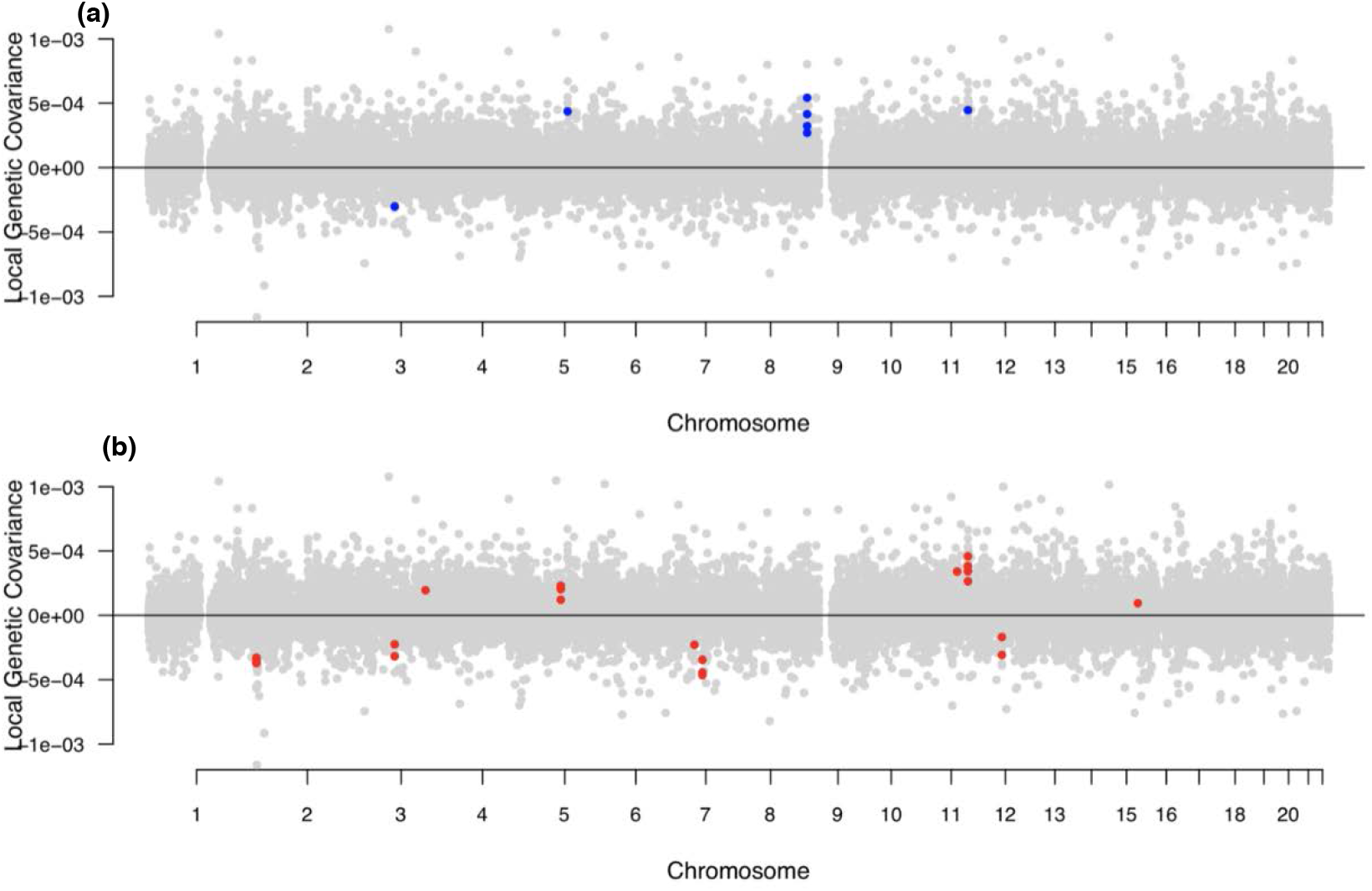
Estimates of local genetic covariance between anxiety traits and PAU traits (a) and anxiety and AC (b) Blue indicates genomic regions with significant local genetic covariances between an anxiety and PAU trait, while red indicates genomic regions with significant local genetic covariances between and anxiety and AC trait (all p < 7.60×10 –7).

While the significant local genetic covariances between PAU and anxiety were generally positive (Table S6, Figure 2a), significant local genetic covariances between AC and anxiety were bidirectional (Table S6, Figure 2b). For example, negative genetic covariances were found at chr2:4 148 784–5 017 and chr7:68 562 932–69 806 895 between anxiety and AC traits (Figures S3-S9). Conversely, we found a locus (chr11:113 105 405–113 958 177) with significant positive covariance between anxiety and AC traits, as well as between anxiety and PAU (Figure S3).

#### Genetic Covariance Stratified by Functional Annotations

Genetic covariance was significantly positive among pairs of anxiety phenotypes and pairs of alcohol use phenotypes for both functional and nonfunctional genomic regions (Figure S10-S11, Table S7). However, between anxiety and alcohol traits, the anxiety meta-analysis and UKB anxiety had significant, positive genetic covariance in the non-functional genome with alcohol dependence and AUDIT-P (p< 0.00089), but not AC. We found no significant tissue-specific genetic covariance between any pairs of anxiety and alcohol use (all p>0.00026), and limited significant genetic covariance within either trait set, for the immune, brain, muscle, gastro-intestinal, epithelium, cardiovascular and other tissues categories (Figures S12-S13, Table S8).

To better understand the shared influence of the brain on these disorders, and because the contribution of genetic variation in genes expressed in specific brain regions is known across many complex traits [14], we used 13 specific brain region annotations to look at genetic covariance of traits for those regions. We found significant, positive genetic covariance across most brain region annotations for pairs of anxiety phenotypes, and for most pairs of alcohol use traits (p< 0.00014; Table S9).

Again, the genetic covariance between anxiety with PAU and AC were distinct (Figures 3–4, S14, Table S9). There was positive genetic covariance of AUDIT-P and the UKB anxiety among loci with expression specific to the amygdala and caudate basal ganglia, and AUDIT-P and the anxiety meta-analysis genetic covariance significantly covaried among loci specifically expressed in the caudate basal ganglia (all p< 0.00014). Additionally, we found significant, positive genetic covariance of alcohol dependence and the anxiety meta-analysis localized to genes with expression specific to the frontal cortex (p< 0.00014) (Figure S14). We found no significant genetic covariance in specific brain regions amongst pairs of anxiety and AC phenotypes. Results of the partitioned heritability and correlation of partitioned heritability analyses also used to assess genetic relationships amongst functional annotations can be found in the Supplementary Results (Tables S10–13 and Figures S15–19).

**Figure 3.**
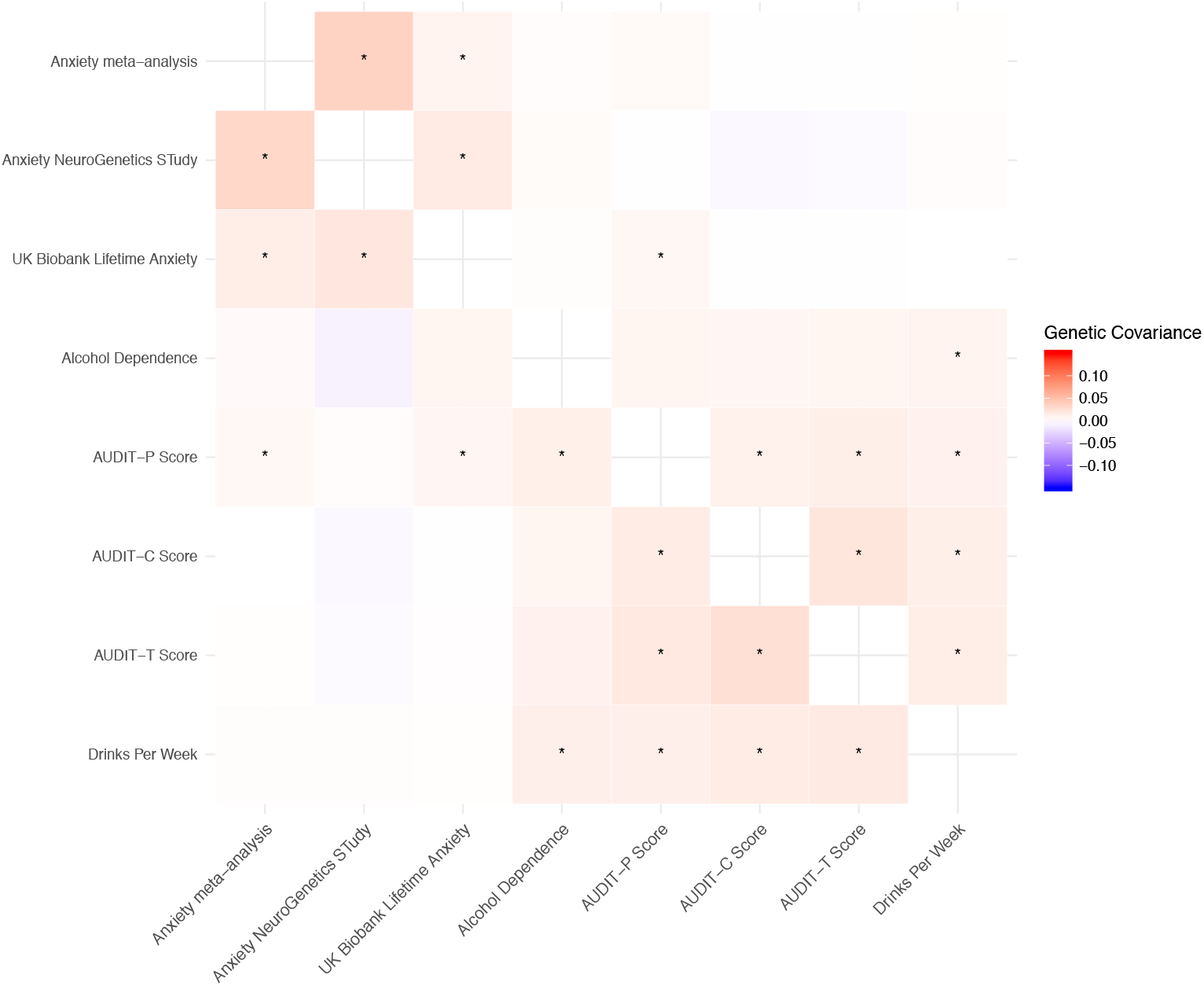
Genetic covariance stratified by brain region: Estimates of genetic covariance in the caudate basal ganglia (lower triangle) and amygdala (upper triangle). Asterisks indicate significant (p< 0.00014) genetic covariance after Bonferonni correction. Results for other specific brain regions can be found in Table S9.

**Figure 4.**
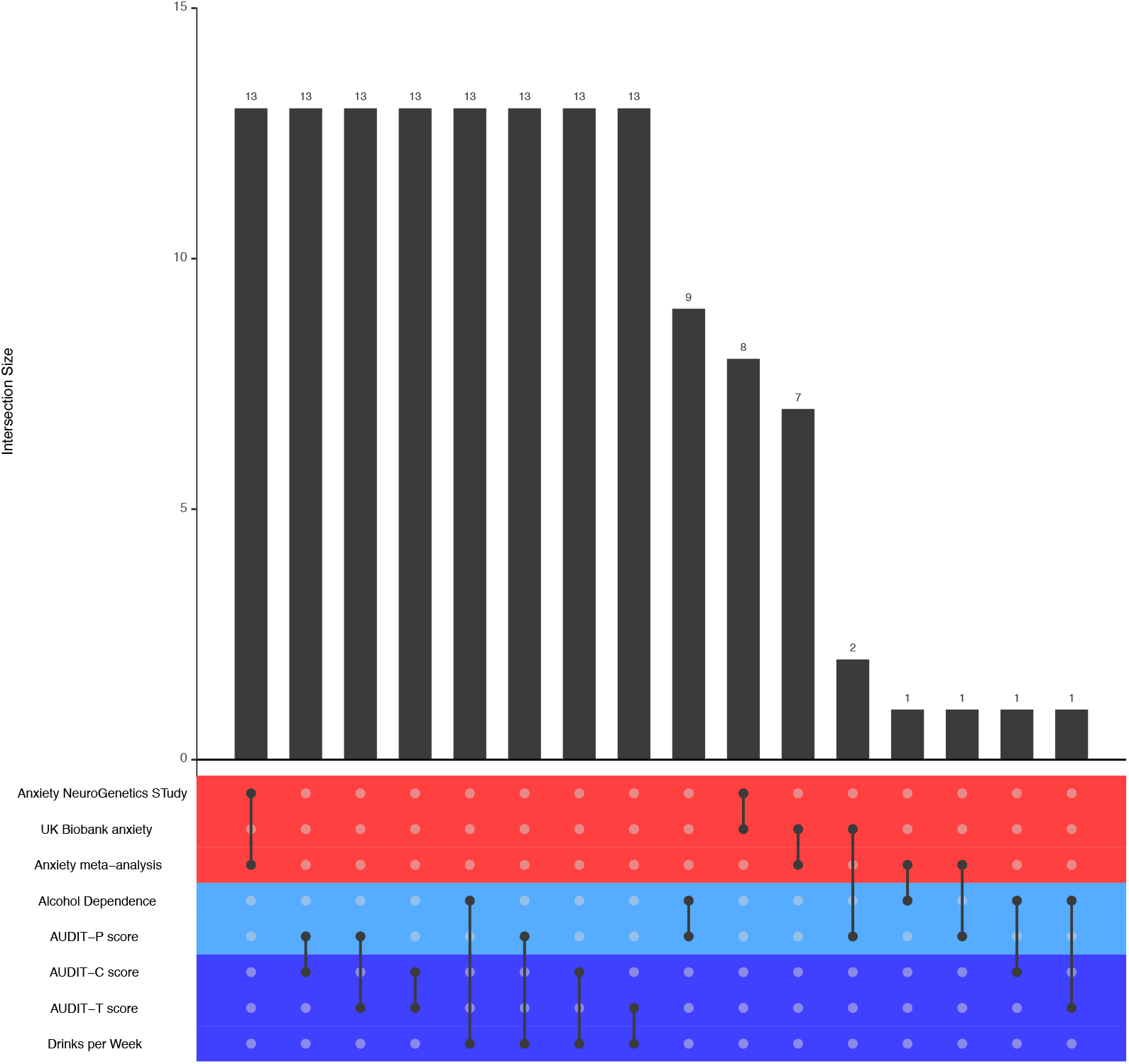
Plot of pairs of traits and the number of shared specific brain region annotations with significant positive genetic covariance (p < 0.00014): Intersect size indicates number of shared annotations. Colors represent the three phenotypic categories of the sets: anxiety disorders (red), PAU (light blue) and alcohol consumption (dark blue).

### Discussion

Our analysis of genome-wide and local genetic covariance unveiled different shared genetic architectures for anxiety phenotypes with PAU compared to AC. While PAU showed a strong, positive genome-wide genetic correlation with anxiety, AC largely did not. At a genome-wide level, this finding supports the hypothesis that alleles which increase genetic risk of anxiety disorders also increase risk of PAU, but not other aspects of alcohol use. That is to say, genetic liability to anxiety disorders may be partly a result of the same genetic components contributing to PAU, but not AC.

Sex-stratified genome-wide genetic correlations also provided insight into the different genetic architectures of these disorders between sexes. Overall, genome-wide genetic correlations of each trait between males and females largely suggested that the genetics underlying these disorders do not differ dramatically between the sexes. We did, however, find that there may be differences between the sexes in terms of the shared genetics of alcohol use and anxiety disorders; specifically, we identified three significant genetic correlations between anxiety and alcohol phenotypes in females, but none in males. Inability to detect significance at the sex-specific level may be a result of the small sample sizes and therefore larger association studies may be required to further explore this hypothesis.

At the scale of individual loci, although we identify 12 specific genomic regions with local genetic correlations between the alcohol use and anxiety traits, we particularly highlight four regions which we consider as possible targets for further functional follow up studies due to the specific genetic makeup of those regions. The locus at chr11:113 105 405–113 958 177, which contains the dopaminergic pathway gene, *DRD2*, showed significant positive covariances between the anxiety meta-analysis and all alcohol use phenotypes except for alcohol dependence, and contains variants associated with worry and neuroticism [27], major depression [28], PAU [12], and AUDIT scores [11]. Researchers also hypothesize that DRD2 expression may substantially moderate stress induced ethanol consumption in mice[29, 30]. Variation in the *DRD2* gene is also associated with connectivity between the basal ganglia and frontal cortices [31], two brain regions identified in the partitioned genetic covariance analysis. In addition to *DRD2*, this region also houses the genes *NCAM1* (Neural Cell Adhesion Molecule 1), *TTC12* (Tetratricopeptide Repeat Domain 12) and *ANKK1* (Ankyrin Repeat And Kinase Domain Containing 1). Together with *DRD2*, these genes make up the NTAD gene cluster [32]. Prior research suggests that this cluster may contribute to various psychiatric disorders as well as the comorbidity of psychiatric disorders [33, 34]. Variants associated with the NTAD gene cluster may therefore represent shared genetic risk factors between anxiety and alcohol use.

Anxiety and alcohol use traits were negatively correlated within a chromosome 3 locus (chr3:85 093 629–86 734 415) which contains *CADM2* (cell adhesion molecule 2), a gene previously found to be associated with AC and various behavioral traits [35], signifying that *CADM2* may influence both of these traits, but in different directions. We also observed negative correlations of ANGST and AC at chromosome 7:68 562 932–69 806 895, which includes *AUTS2* (activator of transcription and developmental regulator). Schumann et al. [36] not only found *AUTS2* to be associated with AC in humans, but also detected expression of the gene in the amygdala and frontal cortex. Additionally, they also found that downregulation of an *AUTS2* homolog in *Drosophila* resulted in reduced alcohol sensitivity, possibly increasing AC [36]. Separately, another study found mice deficient in *Auts2* exhibited a decrease in anxiety-related behaviors [37]. Our results align with these findings, supplementing the idea that *AUTS2* has opposite effects on anxiety disorders and AC. Interestingly, we also observed a significant negative covariance between alcohol dependence and DrnkWk in this region, possibly representing a distinction in the shared genetics of anxiety disorders with the two subgroups of alcohol use traits at this locus. Finally, we isolated a previously unidentified locus on chromosome 9 (10 879 188–11 616 822), with multiple significant positive covariances between anxiety and PAU, but not AC. This locus was associated with worry and neuroticism [27], depression [28] and anxiety [17] in previous studies, but to our knowledge has not been previously implicated in alcohol use.

Analyses of partitioned genetic covariance demonstrated further evidence of distinct relationships of anxiety with PAU and AC. Our analyses revealed unique connections between anxiety disorders and PAU traits across several functional annotations, but notably, this covariance between alcohol use disorders and anxiety disorders is not concentrated across most tissues. Rather, the genetic covariance between anxiety traits and either PAU or AC appears to be distinct among individual brain regions, and highlights the amygdala, caudate basal ganglia and frontal cortex as regions where genetic influences on both anxiety and PAU are manifest. These results align with previous findings using fMRI suggesting that the amygdala, frontal cortex and caudate basal ganglia are associated with anxiety and alcohol use [38–47]. Interestingly, the caudate and frontal cortex are connected extensively, acting together as part of the frontostriatal circuit [48]. This circuit is involved in mood disorders such as major depressive disorder [49] and bipolar disorder [50] and in addiction [51, 52]. Our results bring to light the possibility of an integrative role of this circuit between anxiety and alcohol use, that could also be unique to PAU.

While GWAS sample size and limited reference panels for expression were previously noted as limitations of this study, we also acknowledge that our analyses do not identify specific mechanisms which contribute to the comorbidity of the two disorders or a causal direction. Future analyses will use other rapidly developing statistical genetics approaches to interpret this genetic correlation and test specific hypotheses of function and causal pathways between anxiety traits and PAU and AC. We also note that the lack of diverse ancestry in available summary statistics limits the generalizability of these findings to European-based samples, and this work should be repeated as more diverse samples become available.

In summary, we have identified underlying shared genetic factors of anxiety disorders and alcohol use, partitioned by PAU and AC, while also establish a framework for examining comorbid relationships at multiple scales, from genome-wide patterns to functional annotations to individual genomic loci. In particular, we localize four specific genomic loci as well as the amygdala, caudate basal ganglia and frontal cortex as brain regions particularly important to the underlying genetic relationship of these traits. Furthermore, the genetic factors shared between anxiety phenotypes and PAU and anxiety phenotypes and AC are separable. Future work will evaluate directionality of causation in the alcohol-anxiety relationship and focus on functional characterization of identified genomic loci.

## Data Availability

Summary statistics used in this study were publicly available and access links can be found in original publications.

## Acknowledgements

We thank Matthew C. Keller, Naomi Wray, and Peter Visscher for helpful discussion. This research has been conducted using the UK Biobank Resource under application number 1665. This work was supported by NIH MH100141 (PI: M.C. Keller), AG046938 (PI: C.A. Reynolds/S.M. Wadsworth), 5T32MH016880–38 (PI: John Hewitt), F32AA027435 (PI: E.C. Johnson), R01 NS086933 (PI: C.A. Hoeffer), T32MH016880 (PI: C.A. Hoeffer), the Linda Crnic Institute, Le Jeune Foundation and Institute for Behavioral Genetics. This work utilized the Summit supercomputer, which is supported by the National Science Foundation (awards ACI-1532235 and ACI-1532236), the University of Colorado Boulder, and Colorado State University. The Summit supercomputer is a joint effort of the University of Colorado Boulder and Colorado State University.

This work was conducted using the summary statistics from the Psychiatric Genomics Consortium’s Substance Use Disorders Working group. The Psychiatric Genomics Consortium’s Substance Use Disorders (PGC-SUD) working group is supported by MH109532 with funding from NIMH and NIDA. We gratefully acknowledge prior support from NIAAA and thank all our contributing investigators and study participants who make this research possible.

## Conflict of Interest

The authors declare that there is no conflict of interest.

Supplementary information is available at MP’s website.

